# Characterizing Highly Fermentable Carbohydrate Foods in the Diets of Children with Disorders of Gut-Brain Interaction and Healthy Children

**DOI:** 10.1101/2024.09.23.24314253

**Authors:** Vishnu Narayana, Jocelyn Chang, Ann R. McMeans, Rona L. Levy, Robert J. Shulman, Bruno P. Chumpitazi

**Author notes:** Corresponding Author: Bruno P. Chumpitazi, (BPC). These authors contributed equally to the work (Vishnu Narayana, Jocelyn Chang). These authors also contributed equally to the work.

## Abstract

**Objectives:** Restricting fermentable oligosaccharides, disaccharides, monosaccharides, and polyols (**FODMAPs**) intake can alleviate symptoms in children with gut-brain interaction disorders (**DGBI**). Due to the restrictive nature of the low FODMAP diet (**LFD**), the less restrictive FODMAP Gentle diet (**FGD**) has been suggested. However, the types of high FODMAP foods and carbohydrates commonly consumed by US children are unknown, as is the impact of the FGD on a typical diet. This project aimed to identify the high FODMAP foods and proportions of FODMAP carbohydrates consumed by children with DGBI and healthy children (**HC**), and to determine which usually ingested FODMAPs would be restricted on the FGD.

**Methods:** Three-day diet records from both HC and children with DGBI were analyzed to assess the type of high FODMAP foods and carbohydrates ingested. Results were compared between the groups. The ingested FODMAPs that would be restricted on the FGD was determined.

**Results:** The number of foods ingested daily was similar between children with DGBI and HC (12.3 ± 4.2 vs 12.9 ± 3.4, respectively); high FODMAP foods comprised most foods eaten in both groups. Children with DGBI (vs HC) ate less high FODMAP foods per day (6.5 ± 2.3 vs 8.7 ± 2.4, P<0.0001, respectively). Fructans were the most consumed FODMAP carbohydrate in both groups and children with DGBI (vs HC) consumed fewer fructans, lactose, fructose, and polyols (all P<0.0001). The top 3 food categories consumed in both groups were wheat-containing foods, dairy, and fruits and 100% fruit juices. In children with DGBI, 80.9% of the high FODMAP foods consumed would be limited on the FGD.

**Discussion:** Children with DGBI consume significantly fewer high FODMAP foods and carbohydrates than HC. In both groups, the top consumed FODMAP carbohydrates are fructans, lactose, and fructose. A FODMAP Gentle diet would restrict a large majority of high FODMAP foods consumed by children with DGBI.

## Introduction

Abdominal pain-related disorders of gut-brain interaction (**DGBI**) such as irritable bowel syndrome are highly prevalent. Contributors to symptoms include visceral hypersensitivity, psychosocial distress, gut dysmotility, abnormal gut immune function, abnormal gut microbiota composition, and diet.(1) Dietary culprits include highly fermentable oligosaccharides, disaccharides, monosaccharides, and polyols (**FODMAPs**) which are carbohydrates that are poorly absorbed in the small intestine.(2) After reaching the colon, FODMAPs are rapidly fermented, producing gas and other metabolites.(2-4) The gas and metabolite production, in addition to osmotically-driven water secretion into the lumen of the colon can contribute to DGBI symptom generation.(3) Restricting FODMAP dietary intake through a low FODMAP diet (**LFD**) may ameliorate pediatric and adult DGBI symptoms, including pain, bloating, flatus, diarrhea, nausea, and heartburn.(5-7)

Unfortunately, there also can be challenges with implementing a LFD, including the need for appropriate educational resources (e.g., dietitian knowledgeable about the LFD) and patient adherence.(8, 9) Some patients have reported cost of the diet to be a barrier to adherence.(10) In addition, given the restrictive nature of a LFD, there is concern for potential adverse effects such as decreases in bacteria associated with health (e.g., *Bifidobacteria*), the potential for unintentional weight loss, and risk of developing disordered eating. Due to these possible negative impacts, some experts do not currently recommend a LFD for children.(11-14) This has led to proposals for diets that restrict fewer fermentable carbohydrates (e.g., the “FODMAP Gentle” diet, **FGD**) in an attempt to be more pediatric-friendly.(14, 15) However, determining potentially effective and less restrictive diets such as the FGD for children is hampered by lack of knowledge related to which high FODMAP foods normally are consumed by children and whether the FODMAP intake of healthy children (**HC**) differs from that of children with DGBI. This information is needed to determine the degree to which a less restrictive diet (e.g., FGD) may impact FODMAP intake. Therefore, we sought to identify the high FODMAP foods and proportions of FODMAP carbohydrates consumed by children with DGBI and HC, and to determine which usually ingested FODMAPs would be restricted on the FGD.

## Materials and methods

### Study Design

The data analyzed in this study were dietary intake from a large trial of children with DGBI (NCT03823742) recruited in the study between February 22, 2019 through May 1, 2024, and the dietary intake of HC from a separate study which recruited subjects between May 1, 2013 and October 29, 2018. The Baylor College of Medicine Institutional Review Board for Human Subjects approved both studies. Written informed consent was obtained from study participants’ parents (or guardians) and assent from study participants. Prior to any kind of intervention, parents of participants in both studies completed a 3-day baseline food record of their child’s habitual diet after receiving detailed information on how to complete the diet records.

### Study Population

Participants were children 7-12 years of age from the greater Houston, Texas metropolitan area. Exclusion criteria for both children with DGBI and HC included: children following specific diet alterations (e.g., LFD); those with eating disorders; any recent weight loss (≥ 5 % of body weight); or organic gastrointestinal or systemic diseases (e.g., celiac disease, Crohn’s disease, cancer). Children with DGBI met pediatric Rome IV criteria for an abdominal pain-related DGBI.(16) HC could not have had any healthcare visits related to abdominal pain.

### Data Collection

Following enrollment, 3-day food records of foods and drinks consumed by the children were completed by the parents. Details included the name of the food, amount eaten, and parts of the food if the food had multiple components (e.g., a sandwich had all components listed). Diet records then were analyzed by two research dietitians well-versed in DGBI and FODMAPs and in providing LFD education. Data were entered into the University of Minnesota Nutrition Data System for Research software version 2019.(17) Both children with DGBI and HC kept 2-week pain and stooling diaries as previously reported to insure they met entry criteria.(18, 19)

### Analysis and Classification of Foods

High FODMAP foods are identified using guidelines derived from the Monash University Low FODMAP dietary food guide and smartphone app.(20) High FODMAP foods are further characterized by the type of FODMAP carbohydrate within the food, the food categories to which the foods belong, and whether the foods contained high fructose corn syrup as a major ingredient. Statistical comparisons between the dietary intake of the two groups were made using both chi-square for categorical data and t-testing for continuous data using IBM SPSS Statistics for Windows (IBM Corp. Released 2023. IBM SPSS Statistics for Windows, Version 29.0.2.0 Armonk, NY: IBM Corp). In addition, based on published guidelines related to the FGD, we determined whether the high FODMAP foods consumed by children with DGBI would have been restricted.(14, 15)

## Results

At the time of analysis, 77 children with DGBI had completed baseline diet records for the ongoing study. From the previous study, data from 64 **HC** were available. Self-reported demographic data were similar between groups (Table 1).

**Table 1.** Demographics of Children with DGBI versus Healthy Children.

|  | **DGBI (n=77)** | **HC (n=64)** | **P-value** |
| --- | --- | --- | --- |
| **Age** (years ± SD) | 10.0 ± 1.7 | 9.7 ± 1.5 | 0.25 |
| **Sex** |  |  | 0.68 |
| Female, n (%) | 46 (59.7) | 36 (56.3) |  |
| Male, n (%) | 31 (40.3) | 28 (43.8) |  |
| **Race, ethnicity**, n (%) |  |  | 0.07 |
| White, non-Hispanic | 22 (28.6) | 32 (50.0) |  |
| White, Hispanic | 16 (20.8) | 14 (21.9) |  |
| Black, non-Hispanic | 23 (29.9) | 10 (15.6) |  |
| Black, Hispanic | 4 (5.2) | 2 (3.1) |  |
| Asian, non-Hispanic | 2 (2.6) | 0 (0.0) |  |
| American Indian/ Hawaiian, non-Hispanic | 0 (0.0) | 0 (0.0) |  |
| American Indian/ Hawaiian, Hispanic | 4 (5.2) | 1 (1.6) |  |
| More than one race, non-Hispanic | 1 (1.3) | 3 (4.7) |  |
| More than one race, Hispanic | 1 (1.3) | 2 (3.1) |  |
| Black, no ethnicity identified | 1 (1.3) | 0 (0.0) |  |
| Hispanic, no race identified | 3 (3.9) | 0 (0.0) |  |

Out of a total of 4734 foods consumed in both groups, 2792 (59.0%) were high FODMAP. Children with DGBI consumed fewer high FODMAP foods daily per child than HC (Table 2). High FODMAP foods comprised 1481 (52.5%) and 1311 (68.6%) of total foods consumed by children with DGBI and HC, respectively.

**Table 2.** Daily Food Consumption and High FODMAP Intake in Children with DGBI versus Healthy Children.

|  | **DGBI**<br>**(n=77)** | **HC**<br>**(n=64)** | **P-value** |
| --- | --- | --- | --- |
| **Total Foods Consumed** | 2822<sup>1</sup> | 1912<sup>1</sup> |  |
| Total Foods per subject per day<br>(mean ± SD) | 12.3 ± 4.2 | 12.9 ± 3.4 | 0.36 |
| High FODMAP Foods per subject per day<br>(mean ± SD) | 6.5 ± 2.3 | 8.7 ± 2.4 | <0.001 |
| **FODMAP Sugar, n (%)<sup>2</sup>** |  |  |  |
| Fructans<sup>3</sup> | 850 (30) | 748 (39) | <0.0001 |
| Lactose<sup>4</sup> | 552 (20) | 527 (28) | <0.0001 |
| Fructose<sup>5</sup> | 412 (15) | 570 (30) | <0.0001 |
| Polyols<sup>6</sup> | 82 (3) | 100 (5) | <0.0001 |
| Galacto-oligosaccharides<sup>7</sup> | 82 (3) | 53 (3) | 0.79 |
| HFCS<sup>8</sup> | 248 (9) | 388 (20) | <0.0001 |
| **Food Category, n (%)<sup>2</sup>** |  |  |  |
| Wheat | 557 (20) | 534 (28) | <0.0001 |
| Dairy | 344 (12) | 342 (18) | <0.0001 |
| Fruits and 100% Fruit Juices | 168 (6) | 194 (10) | <0.0001 |
| Beverages (e.g., sodas, juice w/ high fructose<br>corn syrup) | 144 (5) | 93 (5) | 0.71 |
| Condiments (e.g., salad dressing, mayonnaise,<br>ketchup, jelly, jam) | 125 (4) | 74 (4) | 0.35 |
| Sweet Snacks (e.g., candy, cookies, brownies) | 125 (4) | 143 (7) | <0.0001 |
| Vegetables | 111 (4) | 80 (4) | 0.67 |
| Savory Snacks (e.g., chips) | 84 (3) | 90 (5) | 0.002 |
| Sweeteners (e.g., fructose, honey, added<br>polyols) | 64 (2) | 35 (2) | 0.30 |
| Legumes | 42 (1) | 16 (1) | 0.06 |
| Nuts, seeds, nut/seed butters | 8 (<1) | 13 (1) | 0.04 |
| **Top High FODMAP Foods Consumed, n (%)<sup>2</sup>** |  |  |  |
| Milk | 146 (5.2) | 121 (6.3) | .09 |
| Sodas<sup>9</sup> | 69 (2.4) | 31 (1.6) | .05 |
| Fructose-sweetened drinks<sup>9</sup> | 67 (2.4) | 45 (2.4) | 0.96 |
| Breaded Animal Protein<sup>10</sup> | 65 (2.3) | 34 (1.8) | 0.22 |
| Fresh fruit | 62 (2.2) | 74 (3.9) | <0.001 |
| Loaf bread<sup>11</sup> | 61 (2.2) | 63 (3.3) | 0.02 |
| Savory condiments<sup>12</sup> | 58 (2.1) | 18 (0.9) | <0.005 |
| 100% Fruit juice | 44 (1.6) | 46 (2.4) | 0.04 |
| Sweet/enriched bread<sup>11</sup> | 43 (1.5) | 51 (2.8) | <0.01 |
| Pizza<sup>10</sup> | 42 (1.5) | 34 (1.8) | 0.44 |
| Mixed pasta<sup>10</sup> | 35 (1.2) | 35 (1.8) | 0.10 |
| Cookies<sup>11</sup> | 34 (1.2) | 35 (1.8) | 0.08 |
| Seasonings<sup>13</sup> | 23 (0.8) | 41 (2.1) | <0.001 |
| Crackers<sup>11</sup> | 20 (0.7) | 40 (2.1) | <0.0001 |
| Fruit snacks | 20 (0.7) | 35 (1.8) | <0.0005 |

<sup>1</sup>Total count of all foods consumed
<sup>2</sup>Percent out of total foods consumed
<sup>3</sup>Fructans are found in wheat, garlic, onion, legumes, and inulin and chicory root which are types of fiber and the main ingredient in fiber supplements or added to baked goods to increase fiber content.
<sup>4</sup>Lactose is found in dairy products and may be added to some food products such as chocolate bars, flavored chips, and baked sweets.
<sup>5</sup>Fructose is found in many fruits and some vegetables such as apples, pears, and asparagus; it is found in honey and agave syrup and may be added as fructose or high fructose corn syrup to provide a sweet flavor in different food products, such as sodas, jams, ketchup, and baked sweets.
<sup>6</sup>Polyols can be found in different fruits such as peaches, plums, and avocado and may be added in different forms (i.e., sorbitol, mannitol, xylitol) to impart a sweet flavor to different food products, most notably sugar free candies and sugar free gums.
<sup>7</sup>Galacto-oligosaccharides can be found in beans and nuts.
<sup>8</sup>High fructose corn syrup; a syrup of fructose and glucose, usually found in a 55%/45% ratio in food products
<sup>9</sup>Contains high fructose corn syrup
<sup>10</sup>Contains wheat, and onion and/or garlic seasoning
<sup>11</sup>Contains wheat and may contain high fructose corn syrup
<sup>12</sup>Contains onion, garlic, and/or high fructose corn syrup
<sup>13</sup>Contains dried or powdered onion and/or garlic added during at home food preparation process. Does not include packaged foods that already contain spices.

Based on the FODMAP carbohydrate, fructan-containing foods were the most numerous in the diet of both groups, followed by foods containing lactose and fructose (Table 2). Individual foods containing more than one type of FODMAP contributed 501 (33.8%) and 638 (48.7%) of the high FODMAP foods consumed by children with DGBI and HC, respectively. Children with DGBI (vs HC) consumed significantly less fructans, lactose, fructose, and polyols but not galactans.

Based on food category, wheat-containing foods were the most prevalent, followed by dairy and fruits/100% fruit juices (Table 2); children with DGBI consumed significantly fewer of these foods compared to HC. Children with DGBI (vs HC) consumed less FODMAP-containing sweet and savory snacks. There was no significant difference in intake of beverages, condiments, vegetables, and sweeteners.

Based on the high FODMAP-containing foods, children with DGBI consumed most frequently milk, sodas, and fructose-sweetened drinks (e.g., fruit-flavored drinks containing high fructose corn syrup) whereas HC consumed milk, fresh fruit, and wheat-containing loaf bread (Table 2). High FODMAP foods containing high fructose corn syrup as a major ingredient were consumed by 16.7% (248) of children with DGBI and 29.6% (388) of HC.

In children with DGBI, 1198 (80.9%) of the consumed high FODMAP foods would be restricted on a FGD. Of the 283 high FODMAP foods permitted on a FGD, 139 (49.1%) contained high fructose corn syrup as a major ingredient. Of these, beverages and condiments (e.g., sodas, jam/jelly, ready-to-drink lemonade, pancake syrup, and flavored drinks) comprised a significant proportion.

## Discussion

Determining the type of high FODMAP foods and FODMAP carbohydrates consumed by children with DGBI and HC children is important for tailoring educational content and aid in the creation of diets that restrict FODMAP intake. We determined that high FODMAP foods comprise a majority of the foods eaten by both children with DGBI and HC. Children with DGBI (vs HC) consumed fewer high FODMAP foods and several FODMAP carbohydrates. We also identified that a large majority of FODMAP foods normally eaten by children with DGBI would be restricted on a FGD.

We hypothesize the less frequent ingestion of high FODMAP foods by children with DGBI may relate to self-restriction (either intentional or due to unconscious behavior) to avoid worsening their GI symptoms. Supporting the former, a recent study of school children reporting bothersome GI symptoms identified an association between high FODMAP foods and worse GI symptoms.(21) Furthermore, children with IBS self-restrict more foods than healthy children.(22) Additional studies are needed to determine the reasons why children with DGBI eat fewer FODMAPs than HC.

To our knowledge, this is the first study to describe FODMAP intake in children with DGBI and HC under normal dietary circumstances. Fructans were the FODMAP carbohydrate ingested most commonly in both groups. A U.S. Department of Agriculture survey from 1994-1996 estimated that 75% of fructan intake in individuals was from wheat and 25% from onions.(23) Our study findings are in agreement with the survey data, as wheat-containing products, particularly breaded meat products such as chicken nuggets and wheat bread, were the primary contributors to fructan intake for both groups. Onion and garlic powders via seasoning contributed to fructan intake to a lesser degree. Some of the consumed foods containing onion and garlic powder seasonings included flavored chips, processed and seasoned meats, and seasoned beans (i.e., refried beans). Fructans, which arrive essentially intact into the colon after consumption, are often a focus of a low FODMAP diet as fructans have been found to induce more DGBI symptoms than other FODMAPs.(24) Given our finding that wheat (a FODMAP) is the most commonly ingested category of food, particular attention to restriction of wheat products should be a focus of a reduced FODMAP diet.

After fructans, lactose and fructose were the most commonly ingested FODMAPs. Lactose malabsorption is not more common in those with DGBI than HC but lactose intolerance appears to be; likely related to visceral hyperalgesia commonly found in those with DGBI.(25) Similar to education regarding wheat ingestion, our data would suggest emphasis also should be given to a reduction in fructose-sweetened drinks including sodas and 100% fruit juice in children with DGBI. In contrast, a global reduction in fresh fruit might not be warranted given its value as an important component of a healthy diet.

We found that the vast majority of high FODMAP foods commonly consumed by the children with DGBI would be restricted on a FGD which has been proposed as an alternative, pediatric-friendly and less restrictive version to the traditional LFD.(14, 15) The United Kingdom’s National Institute of Clinical Excellence (NICE) dietary guidelines that reduce FODMAP intake also has been recommended but data are conflicting as to whether it has similar (worse or better) efficacy as the traditional LFD in adults and children.(14) One particular advantage of the FGD may be its relative ease related to education and adherence as only 15 different food and/or food types (e.g., wheat) are restricted. However, its efficacy in children with DGBI remains to be determined.

Of the high FODMAP foods consumed by the children in our study that would be allowed on a FGD, about half contained high fructose corn syrup as a major ingredient-not surprising given its prevalence in many foods in the US. Important to note is that the FGD was developed in Australia where high fructose corn syrup is not added to food as frequently as in the US.(15) Thus, in the US, restriction of foods containing this FODMAP (e.g., sodas, jams) is likely needed when children are placed on a FGD.

There are some limitations to our study. Parents may not have accurately recorded food intake of their children with the use of food records. However, 3-day food records are considered a valid method to record food consumption and are considered a better method of food capture compared to other approaches such as 24-hour recall.(26) Children were recruited from a single geographic area and diets may differ based on geographic location. On the other hand, our study population included a racially and ethnically diverse population which may increase the generalizability of our findings. Further studies in different geographic areas can be conducted to confirm our findings

A major strength of our study is that, to our knowledge, it is the first to characterize and compare the FODMAP intake of children with DGBI and HC. Also, the two populations were well-matched in terms of demographics, increasing the rigor of the findings. Additionally, the dietitians participating in the study had experience with analyzing food records for high FODMAP foods and maintained high quality in their review, including follow-up with families, if needed, regarding specific food record entries.

In summary, both children with DGBI and HC consume a significant amount of high FODMAP foods, though children with DGBI consume relatively less.. Fructans, lactose, and fructose were the most consumed FODMAP carbohydrates via wheat-containing foods, dairy, and fruits/100% fruit juices. Targeted and less restrictive diets such as the FGD, with modification based on our findings, may effectively restrict a large majority of the high FODMAP foods consumed by children with DGBI.

## Data Availability

That data may be found in the submission.

